# Immunogenicity of the third and fourth BNT162b2 mRNA COVID-19 boosters and factors associated with immune response in systemic lupus erythematosus and rheumatoid arthritis patients

**DOI:** 10.1101/2022.03.15.22272350

**Authors:** Theerada Assawasaksakul, Seelwan Sathitratanacheewin, Preeyaporn Vichaiwattana, Nasamon Wanlapakorn, Yong Poovorawan, Yingyos Avihingsanon, Nawaporn Assawasaksakul, Wonngarm Kittanamongkolchai

## Abstract

**Objectives:** To evaluate the safety and immunogenicity of third and fourth BNT162b2 boosters in systemic lupus erythematosus (SLE) and rheumatoid arthritis (RA) patients.

**Methods:** SLE and RA patients aged 18-65 years who completed a series of inactivated, adenoviral vector, or heterogenous adenoviral vector/mRNA vaccines for at least 28 days were enrolled. Immunogenicity assessment was done before and day 15 after each booster vaccination. The third BNT162b2 booster was administered on day 1. Patients with suboptimal humoral response to the third booster dose (anti-receptor binding domain (RBD) IgG on day 15 < 2,360 BAU/mL) were given a fourth BNT162b2 booster on day 22.

**Results:** Seventy one SLE and 29 RA patients were enrolled. The third booster raised anti-RBD IgG by 15 fold and patients with positive neutralizing activity against the Omicron variant increased from 0% to 42%. Patients with positive cellular immune response also increased from 55% to 94%. High immunosuppressive load and initial inactivated vaccine were associated with lower anti-RBD IgG titer.

Fifty four patients had suboptimal humoral responses to the third booster and 28 received a fourth booster dose. Although anti-RBD IgG increased further by 7 fold, no significant change in neutralizing activity against the Omicron variant was observed. There were 2 severe SLE flares that occurred shortly after the fourth booster dose.

**Conclusions:** The third BNT162b2 booster significantly improved humoral and cellular immunogenicity in SLE and RA patients. The benefit of a short interval fourth booster in patients with suboptimal humoral response was unclear.

**Key messages:** *What is already known about this subject?:*

- The SARS-CoV-2 omicron variant (B.1.1.159) has multiple mutations that have resulted in greater escape from immune protection elicited by COVID-19 vaccines.
- More attenuated immune response to SARS-CoV-2 vaccination has been observed in patients with autoimmune rheumatic diseases. The additional third dose of SARS-CoV-2 vaccine has been recommended in immunocompromised populations.
- Some immunocompromised patients have a suboptimal humoral response to a third booster dose. Factors associated with poor immune response have not been adequately studied.
- Administration of more than 3 doses has been shown to enhance immune response in some severely immunocompromised patients.

*What does this study add?:*

- The third BNT162b2 booster was well tolerated, and significantly improved both humoral and cellular immunogenicity in SLE and RA patients previously vaccinated with either inactivated, adenoviral vector, or heterogenous adenoviral vector/mRNA vaccines.
- High intensity of immunosuppressive therapy and initial inactivated vaccine were associated with lower humoral immune response to the third BNT162b2 booster.
- Administration of a fourth BNT162b2 booster in poor humoral immune responders may not offer additional protection against the omicron variant, and flares were observed in SLE patients.

*How might this impact on clinical practice or future developments?:*

- This study supported a third BNT162b2 booster dose administration in SLE and RA patients to enhance immune protection against the Omicron variant.
- Patients who receive a high dose of immunosuppressive therapy or initial inactivated vaccine could be unprotected from SARS-CoV-2 infection. Benefits and risks of additional boosters or second generation of SARS-CoV-2 vaccine should be further studied.

## Introduction

The newly emerged B.1.1.159 (Omicron) variant of severe acute respiratory syndrome coronavirus 2 (SARS-CoV-2) has become the dominant strain globally. Its multiple mutations have resulted in greater escape from immune protection elicited by COVID-19 vaccines^1^. Recent studies in healthy populations suggested that the third booster dose of mRNA vaccine enhanced protection against the Omicron variant, although neutralizing antibody titers were reduced by 7-fold compared to the ancestral variant ^2-4^.

In patients with autoimmune rheumatic diseases, a more attenuated immune response to SARS-CoV-2 vaccination has been observed^5^. According to a recent study, a third dose of mRNA 1273 vaccine significantly increased anti-receptor binding domain (RBD) titer in transplant recipients compared to placebo. However, in approximately half of the patients who received a booster, the anti-RBD titer remained low (<100 U/mL)^6^. Clinical factors associated with the immunogenicity in immunosuppressed populations have not been adequately studied. Several booster doses may be required to achieve adequate immune protection^7^. The safety and risk of disease flare associated with repetitive vaccination in patients with autoimmune rheumatic diseases remain unknown.

We aimed to evaluate safety and immunogenicity of the third and fourth BNT162b2 booster doses in systemic lupus erythematosus (SLE) and rheumatoid arthritis (RA) patients previously vaccinated with either adenoviral vector, inactivated or heterogenous adenoviral vector/mRNA regimens. Factors associated with humoral immune response after the third booster dose were also evaluated.

## Methods

### Trial design

This is a prospective, single arm, open-labeled study, investigating the safety and immunogenicity of third and fourth mRNA vaccine (BNT162b2, Pfizer–BioNTech) booster doses in SLE and RA patients who have previously been vaccinated with inactivated SARS-CoV-2 COVID-19 vaccine (CoronaVac or Sinopharm), adenovirus-vectored vaccine (AZD1222, AstraZeneca), or heterologous regimen of AZD1222/BNT162b2. This trial was reviewed and approved by the Institutional Review Board of the Faculty of Medicine, Chulalongkorn University (1425/2021) and registered in the Thai Clinical Trial Registry (TCTR20211220004).

### Participants

Eligible patients were aged 18 to 65 years who met the classification criteria for systemic lupus erythematosus or rheumatoid arthritis and had completed a series of inactivated vaccine (CoronaVac or Sinopharm), AZD1222 or AZD1222/BNT162b2 for at least 28 days. Intervals between the first and second dose of COVID-19 vaccines were stipulated by vaccine types: 4 weeks for inactivated vaccine, 8-12 weeks for AZD1222/AZD1222, and 4-8 weeks for AZD1222/BNT162b2. Exclusion criteria were history of SARS-CoV-2 infection, allergy to a vaccine component, pregnancy, breastfeeding, and active disease at enrollment. Recruitment occurred at the rheumatology and nephrology clinic in a tertiary referral center in Bangkok, Thailand. Participants meeting eligibility criteria were invited to participate in the study and provided written informed consent.

### Procedures

At baseline, demographic information, current medications, disease activity, and blood samples for immunogenicity analyses were collected. On day one, 30 ug of BNT162b2 was administered intramuscularly via the deltoid region. On day 15, blood samples were collected in order to assess immune responses. On day 22, those with suboptimal humoral response, defined as anti-RBD IgG less than 2,360 BAU/mL (16,503 AU/mL) after the third booster, were given the fourth BNT162b2 booster at 30 ug. Blood sample collection for immune responses assessment was performed again on day 36 (Figure 1).

**Figure 1:**
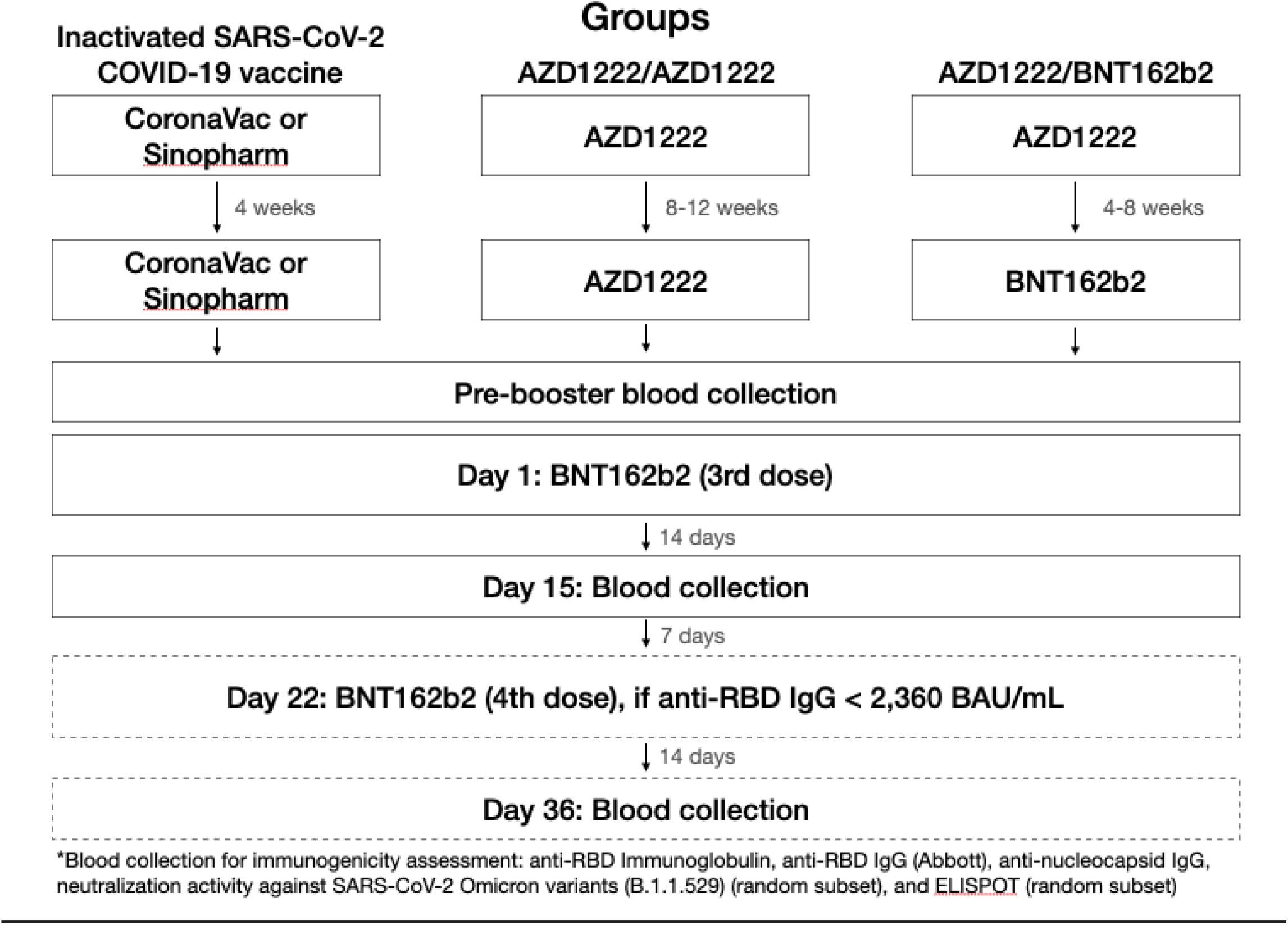
Schematic diagram of the study.

We chose the anti-RBD IgG (Abbott Diagnostics) cut-off value of 2,360 BAU/mL (16,503 AU/mL) as it corresponded to vaccine efficacy of 90% for wild-type^8^. We aimed for high vaccine efficacy due to the spread of vaccine-resistant variants including Delta (B.1.617.2) and Omicron (B.1.1.529) variants.

### Immunogenicity assessment

The humoral responses were measured by serum binding IgG levels against RBD of the SARS-CoV-2 spike protein using the SARS-CoV-2 IgG II Quant assay (Abbott Diagnostics, Abbott Park, Ill). Values of 50 arbitrary U/mL (AU/mL) were considered positive. The numerical AU/mL is converted to binding antibody units per milliliter (BAU/mL) by multiplying it by 0.142. The SARS-CoV-2 cPassTM surrogate virus-neutralizing test (GenScript, Piscataway, NJ) was used to evaluate the neutralizing activity against the SARS-CoV-2 Omicron variant in a subset of pre and post-booster samples. Inhibition levels greater than 30% were considered positive. The SARS-CoV-2 IgG assay (Abbott Diagnostics, Abbott Park, Ill) was used to quantify the nucleocapsid protein of SARS-CoV-2 to evaluate for recent SARS-CoV-2 infection. Positive values were defined as 1.4. The cellular immune responses were measured by human IFNγ-ELISpot assay (ELISpot) in a random subset of patients. Positive responses were defined as >50 spots per 10_ peripheral blood mononuclear cells (PBMC).

### Immunosuppressive treatments

Due to various immunosuppressive treatment regimens used in our cohort, we adopted and modified the Vasudev score^9^ to calculate the overall immunosuppressive load. One unit of immunosuppression was assigned for each of the following doses of immunosuppressive medications: prednisone 5 mg/day, mycophenolate mofetil(MMF) 500 mg/day, azathioprine 100 mg/day, cyclosporine 100 mg/day, tacrolimus 2 mg/day, leflunomide 10 mg/day and methotrexate 15 mg/week(**Table 1**). The average dose of immunosuppressive medication during the first 30 days were used for immunosuppressive unit scale calculation. Adjustment of immunosuppressive medications during such periods were not allowed unless clinically indicated.

**Table 1:** Immunosuppressive load calculation scale modified from Vasudev score (9). One unit of immunosuppression was assigned to the corresponding doses of agents.

| Immunosuppressive | Unit dose | Immunosuppression unit |
| --- | --- | --- |

| medication |  |  |
| --- | --- | --- |
| Prednisolone | 5 mg/day | 1 |
| Azathioprine | 100 mg/day | 1 |
| Cyclosporine | 100 mg/day | 1 |
| Tacrolimus | 2 mg/day | 1 |
| Mycophenolate mofetil | 500 mg/day | 1 |
| Methotrexate | 15 mg/week | 1 |
| Leflunomide | 10 mg/day | 1 |

### Safety and reactogenicity assessment

Thanou modified SELENA-SLEDAI Flare Index was used to determine SLE flares. Disease activity of RA patients was measured by Disease Activity Score 28 ESR (DAS28-ESR) in a subset of patients. The local and systemic reactogenicity were reviewed on day 15, and reviewed again on day 36 in those who received the second booster.

### Statistical analyses

Continuous variables were expressed as mean (standard deviation) or mean (95% confidence interval (CI)). Antibody levels were presented as geometric mean titers (GMT) with 95% CI, or median with range when appropriate. Geometric means were computed by natural log transformation of the data points and calculating the mean and 95% confidence interval on the transformed data. The ln-transformed mean and 95% confidence interval were transformed back to the original scale.

The Student’s t-test was used to compare the differences between two groups. Multiple groups were compared using one-way ANOVA, followed by the Bonferroni post hoc test. Linear regression was used to test for associations.

Univariable and multivariable linear regression was performed to determine the relationship between clinical factors and immune responses after the third booster dose. P<0.05 was considered statistically significant. Statistical analysis was performed using JMP software V.13.2.1 (SAS Institute, Cary, North Carolina, USA) and dot-plot graphs were created using GraphPad Prism V.4.03 (GraphPad Software, La Jolla, CA, USA).

## Results

One hundred patients were enrolled with 71 having SLE and 29 having RA. All of the patients had stable disease. Ninety three percent of patients were women. The majority of SLE patients received MMF and the majority of RA patients received methotrexate. (**Table 2**)

**Table 2:**
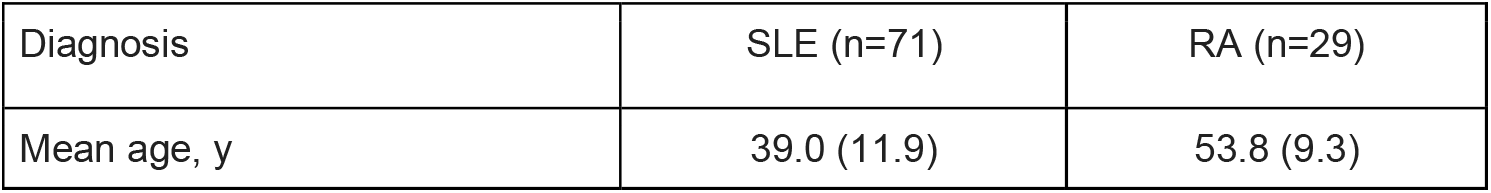
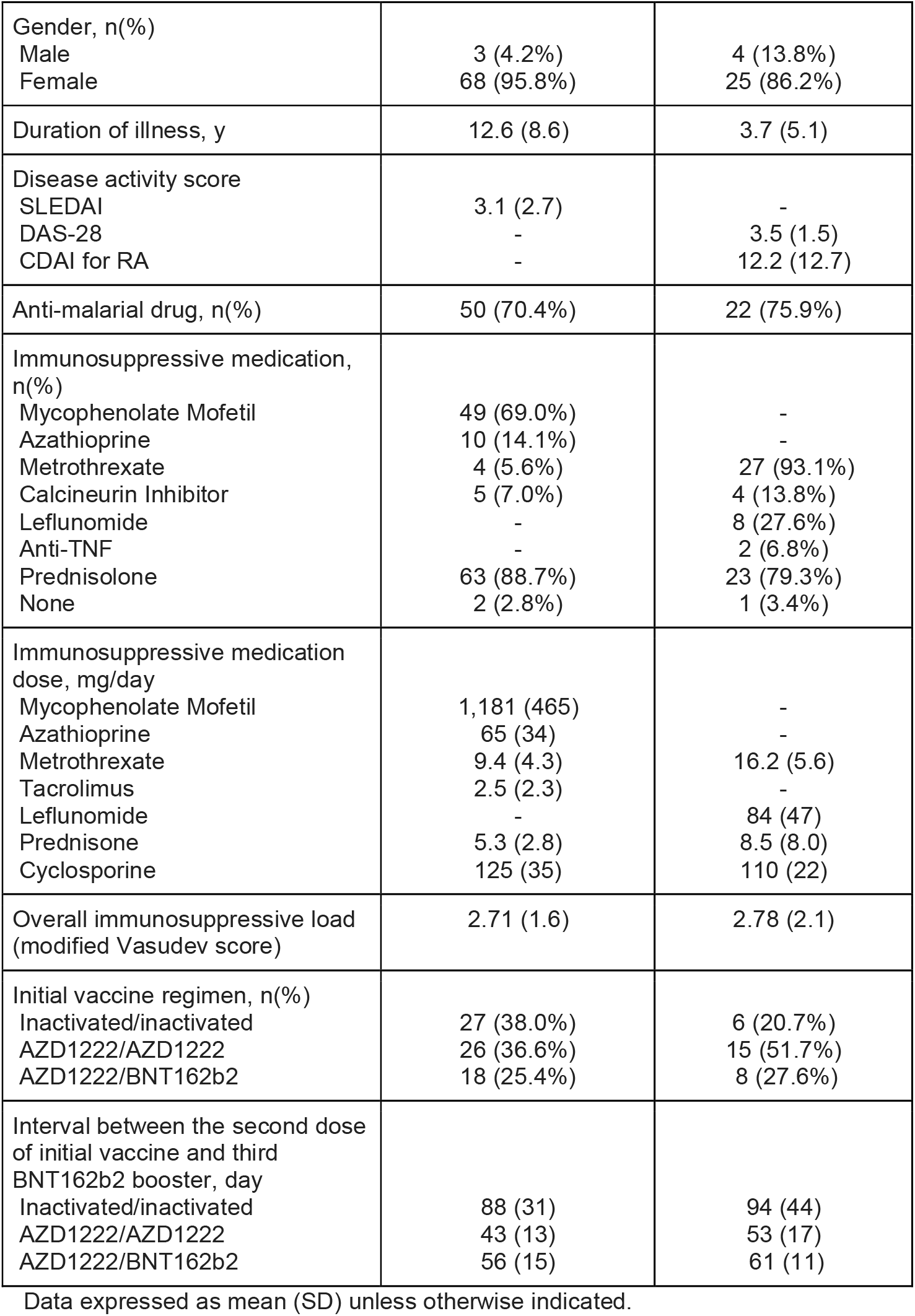
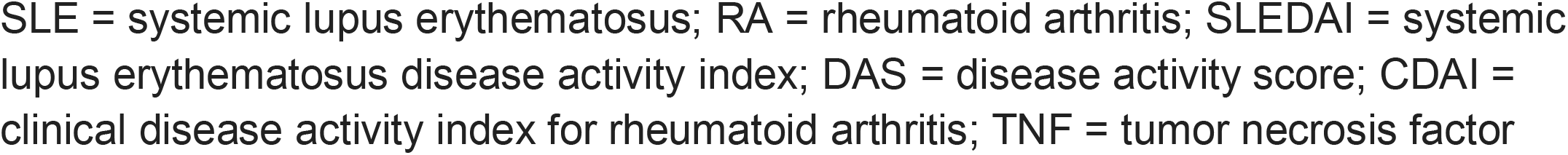
Demographics and clinical characteristics of study participants

Because ADZ1222 was primarily administered to the elderly in Thailand, it was given to slightly over half of the RA patients (51.7%) who tended to be relatively older. Vaccine administration was more equally divided among SLE patients with 38% receiving inactivated vaccines, 36.6% receiving ADZ1222/ADZ1222 and 25.4% receiving AZD1222/BNT162b2. The inactivated vaccine had the longest interval between the second and third booster doses because it was the first vaccine series available in Thailand. (**Table 2**)

### Humoral immune responses following the third and fourth BNT162b2 booster doses

#### Anti-RBD IgG

Anti-nucleocapsid antibodies were undetectable in all pre and post-booster serum samples indicating that no covid-19 infection occurred during the peri-booster period. After the third booster, the GMT of anti-RBD IgG increased by 15 fold (from 69 (95% CI, 42 to 112) BAU/mL to 1034 BAU/mL (95% CI, 677 to 1577), p<0.0001) (**Figure 2a**). Patients with anti-RBD IgG levels of more than 2,360 BAU/mL increased from 2% to 46% (p<0.0001)

**Figure 2:**
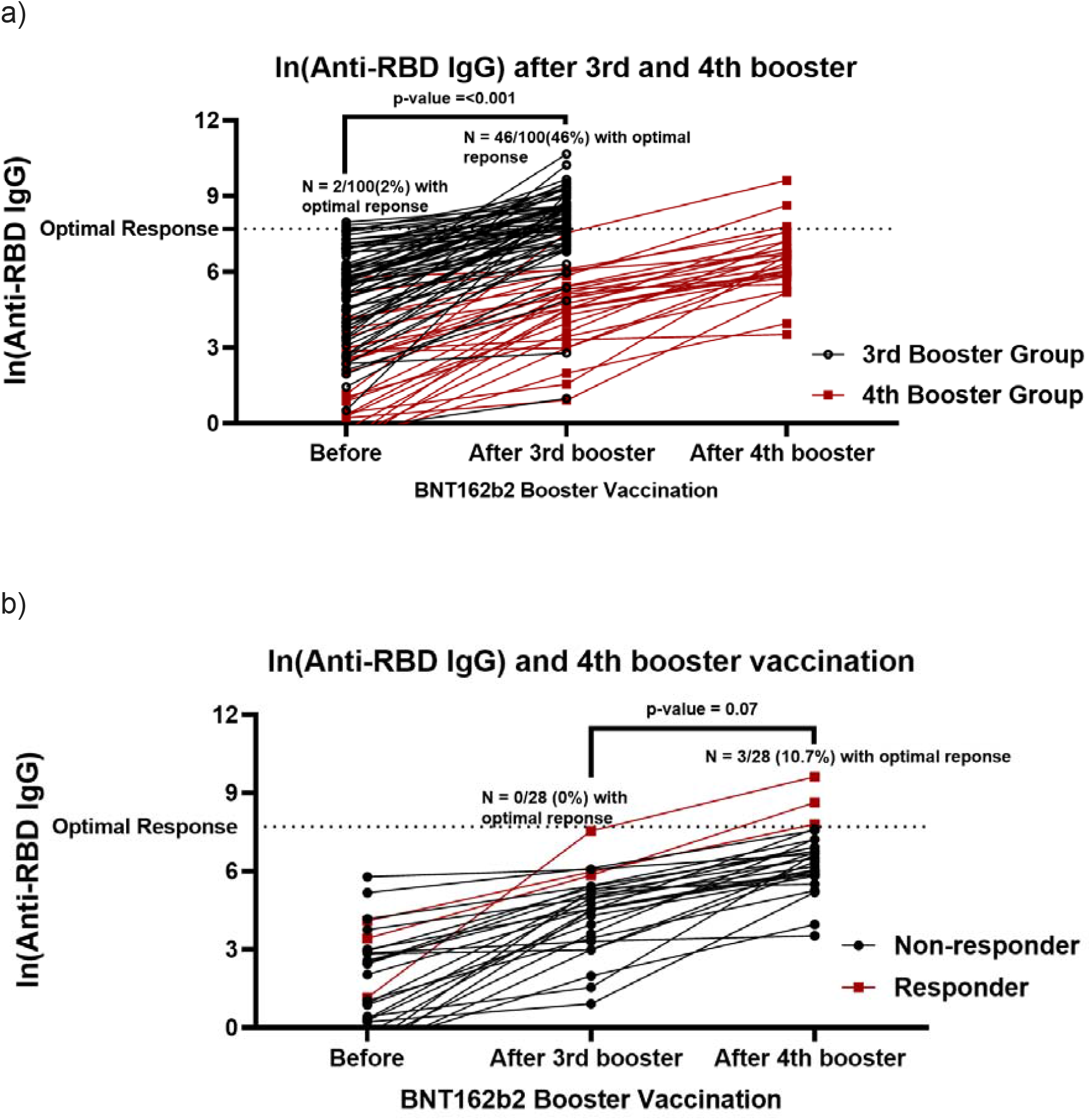
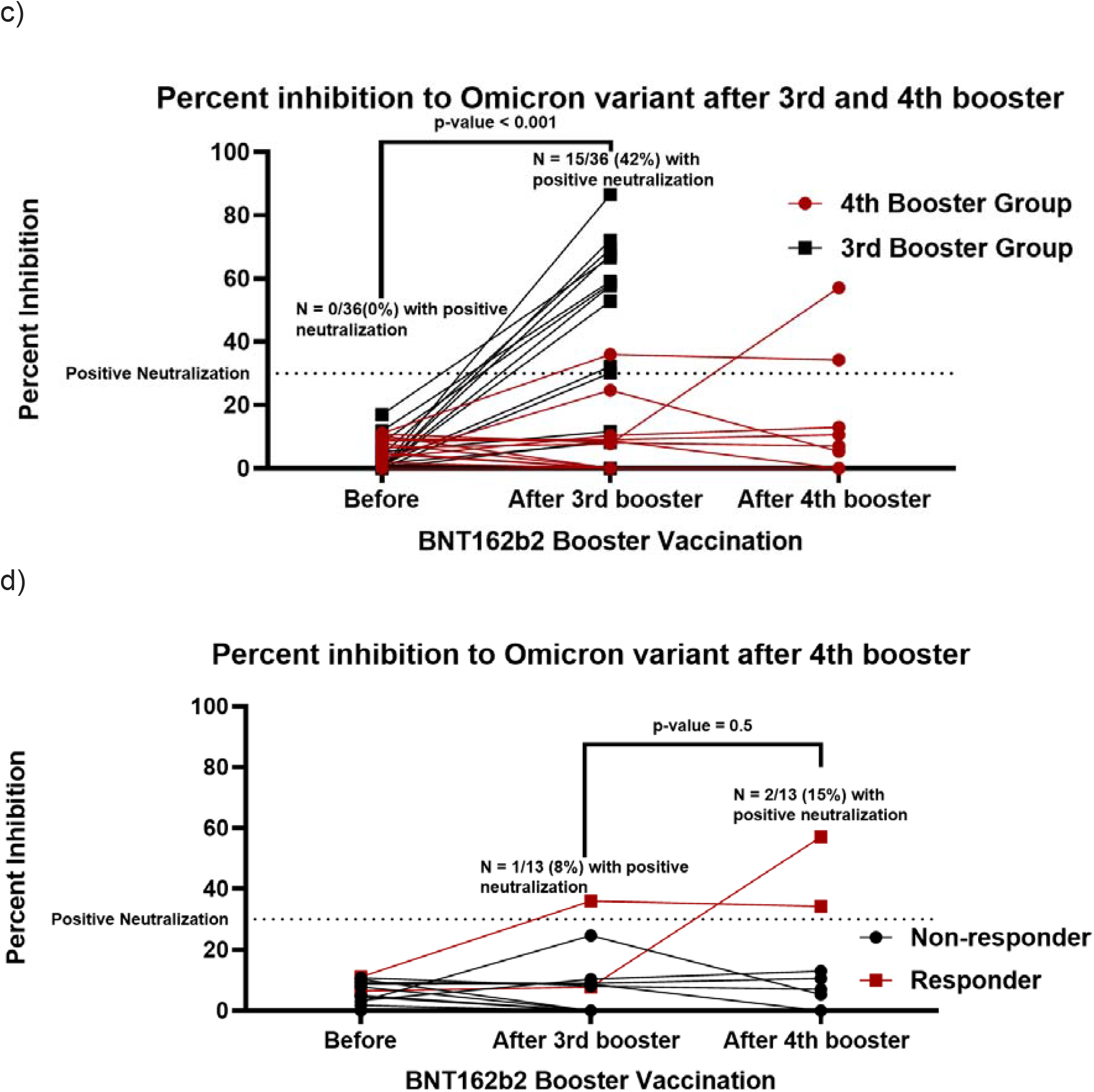
Humoral immune response before and after the third and fourth BNT162b2 booster.Optimal humoral response defined as anti-RBD IgG more than 2,360 BAU/mL (16,503 AU/mL) after the third booster.

Of 54 patients with suboptimal responses to the third booster vaccine (anti-RBD IgG titer < 2360 BAU/mL on day 15), 28 received the fourth BNT162b2 booster on day 22. The remaining patients refused to have another booster dose. The GMT of anti-RBD Ig increased by 7 fold after the fourth booster (88 BAU/mL (range 49-155) to 644 BAU/mL (range 398-1,041), p<0.0001), and 3 patients (11%) had anti-RBD IgG > 2,360 BAU/mL. (**Figure 2b**)

#### Neutralizing activity against Omicron variant

We randomly selected 12 SLE patients from each initial vaccine group (inactivated SARs-CoV2 vaccine, AZD1222/AZD1222, and AZD1222/BNT162b2) to assess for neutralizing activity against the Omicron variant (n = 36). The level of anti-RBD IgG corresponded with the neutralizing activity (r-square = 0.42, p < 0.0001).

Before receiving a third booster dose, none of the patients demonstrated positive neutralizing activity against the Omicron variant. After the third booster dose, 15 patients (42%) had positive neutralizing activity (>30% inhibition), and the mean percent inhibition increased from 5.1% (95% CI, 3.3% to 6.8%) to 28.0 % (95% CI, 17.7% to 38.4%) (p<0.0001) (**Figure 2c**).

Of 36 patients, 13 were subsequently given the fourth booster dose according to the protocol. The neutralizing activity after the third and fourth booster dose was not statistically different (p=0.7). There were only 1 (7.7%) and 2 (15.4%) of the 13 patients who demonstrated positive neutralizing activity against the Omicron variant after the third and fourth booster dose, respectively (**Figure 2d**).

#### Factors associated with humoral immune response

The initial inactivated vaccine and high immunosuppressive load were associated with low anti-RBD IgG titer, even adjusting for other covariates including age, sex, diagnosis, and vaccine interval between the second and the third booster dose (**Table 3**). **Figures 3a,b,c** illustrate the relationship between total immunosuppressive load and type of initial vaccine with anti-RBD IgG levels.

**Table 3:** Association between clinical factors and ln(anti-RBD IgG) at day 15 following a third BNT162b2 booster in SLE and RA patients.

|  | Univariable analysis |  |  | Multivariable analysis |  |  |
| --- | --- | --- | --- | --- | --- | --- |
| Characteristics | Regression coefficient | 95% CI | P-value | Regression coefficient | 95% CI | P-value |
| Age | 0.02 | -0.01, 0.05 | 0.2 | -0.01 | -0.05, 0.03 | 0.55 |
| Sex<br>Male<br>Female | Ref<br>-0.18 | -0.44, 0.66 | 0.7 | Ref<br>-0.12 | -0.91, 0.66 | 0.75 |
| Diagnosis<br>SLE<br>RA | Ref<br>0.30 | -0.16, 0.77 | 0.2 | Ref<br>0.30 | -0.22, 0.83 | 0.25 |
| Initial vaccine regimen<br>AZD1222/BNT162b2<br>AZD1222/AZD1222<br>**Inactivated/Inactivated** | Ref<br>0.02<br>**-0.71** | -0.54, 0.58<br>**-1.31, -0.12** | 0.95<br>**0.02** | Ref<br>0.06<br>**-0.85** | -0.62, 0.75<br>**-1.65, -0.05** | 0.85<br>**0.03** |
| Interval between the second dose of initial vaccine and third BNT162b2 booster | 0.002 | -0.01, 0.02 | 0.8 | 0.01 | -0.01, 0.03 | 0.27 |
| **Total immunosuppressive load** | **-0.41** | **-0.64, -0.18** | **0.0007** | **-0.39** | **-0.63, -0.14** | **0.002** |

SLE = systemic lupus erythematosus; RA = rheumatoid arthritis
Multivariable analysis

**Figure 3a:**
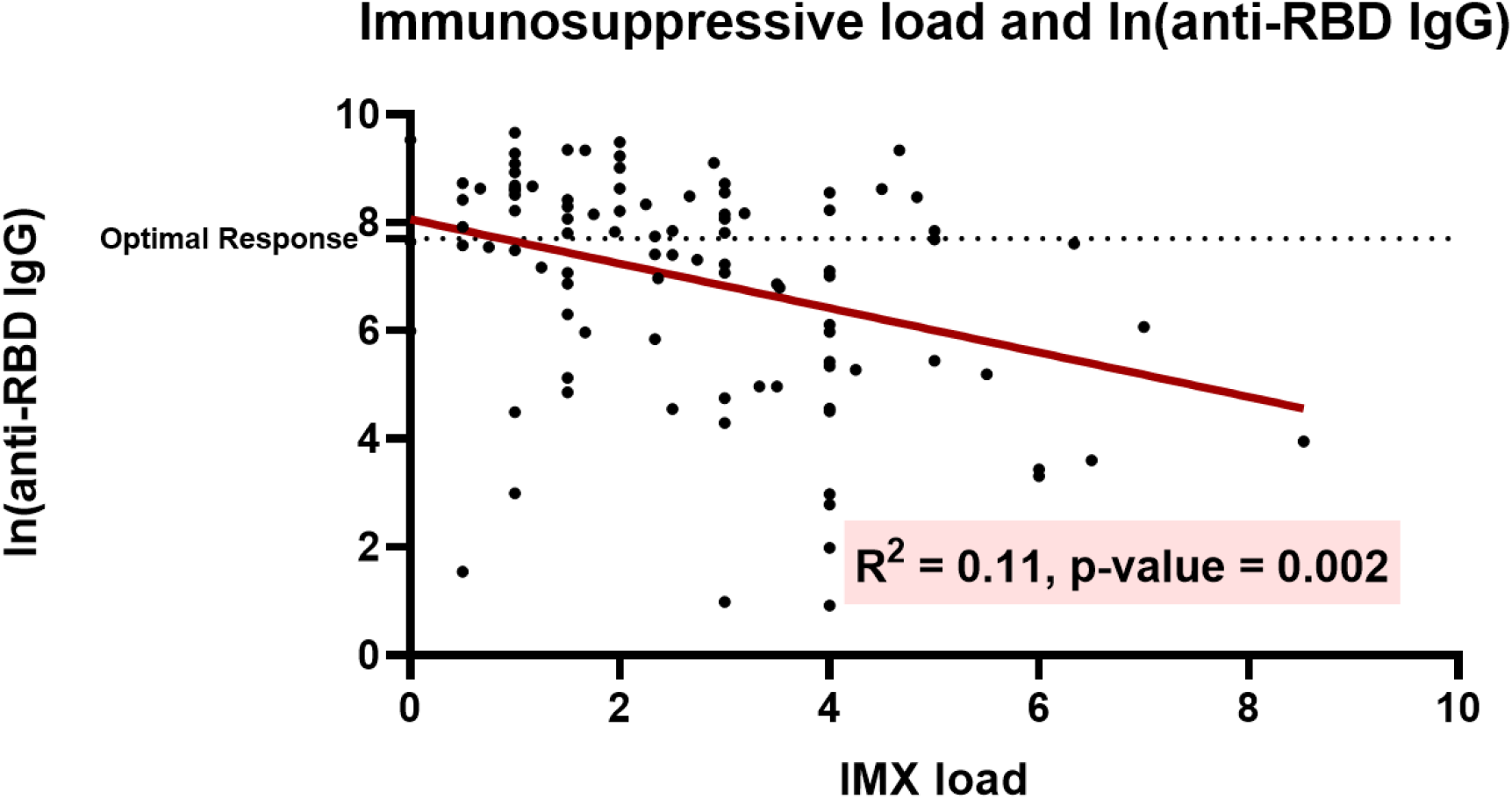
Relationship between total immunosuppressive load and anti-RBD IgG level after the third booster dose.

**Figure 3b:**
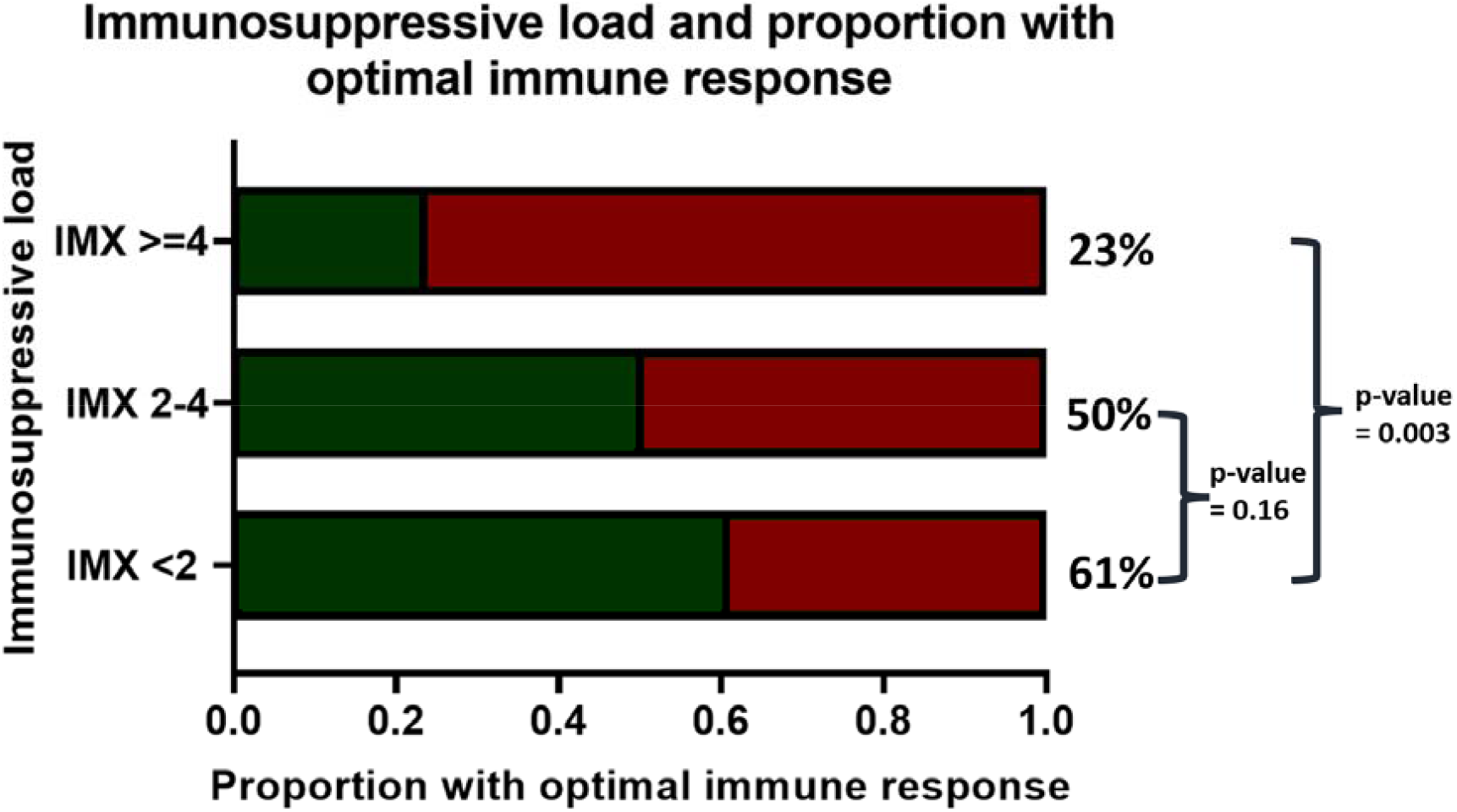
Proportion of patients with SARS-CoV-2 anti-spike receptor binding domain IgG antibody > 2360 BAU/mL by immunosuppressive load (0 to <2, 2 to <4 and 4 or more).

**Figure 3c:**
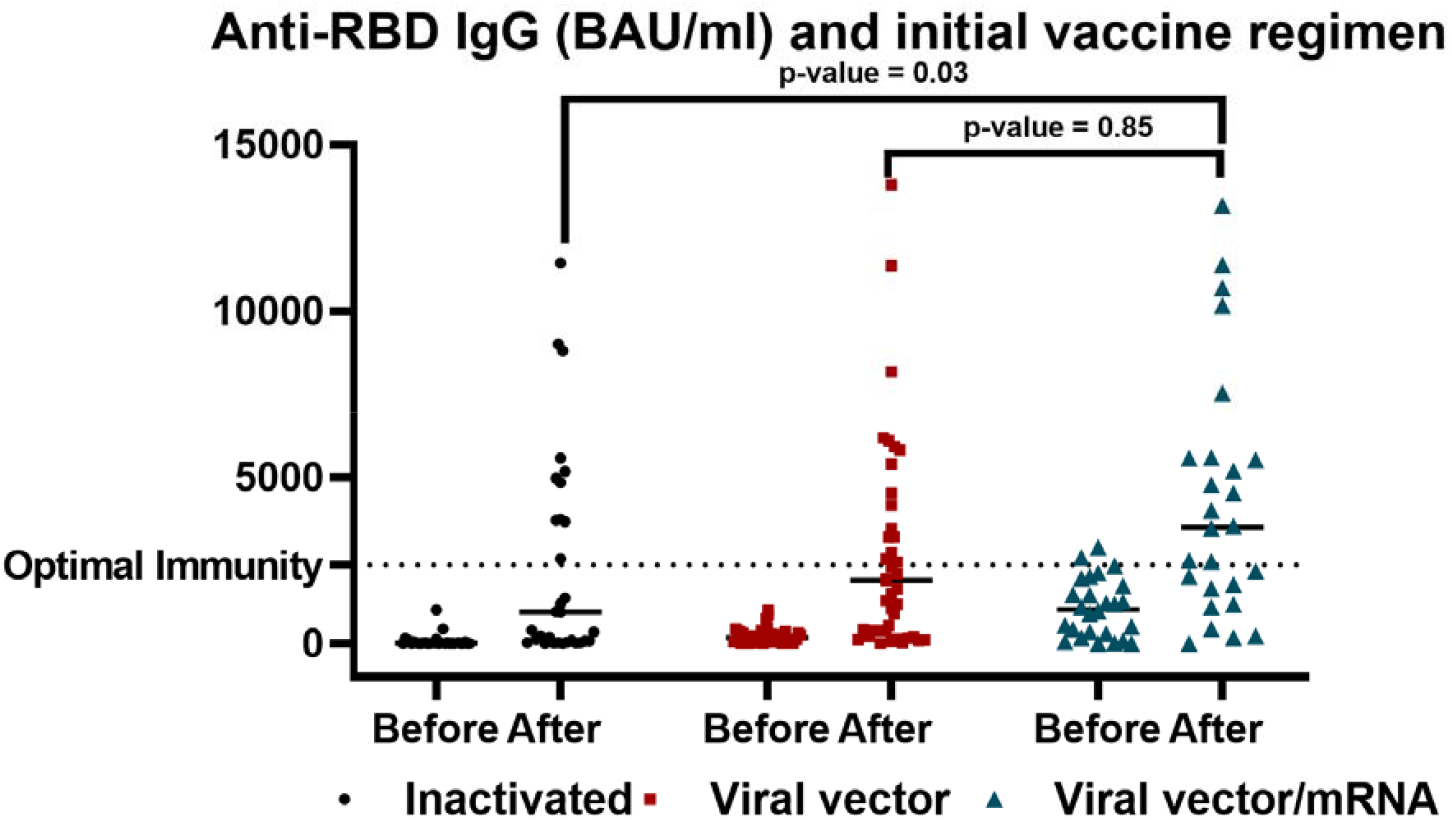
SARS-CoV-2 anti-spike receptor binding domain IgG antibody before and after the third BNT162b2 booster dose by initial vaccine types.

### Cellular immune responses

The cellular immune response of 47 randomly selected patients was assessed before and 14 days after the third booster dose. Twenty one, 15 and 11 patients received inactivated vaccines, AZD1222/AZD1222 and AZD1222/BNT162b2 as initial vaccines, respectively. Thirty four patients (72%) had SLE and the rest had RA.

After a third booster, the proportion of individuals who had a positive ELISPOT test increased from 55% to 94%. Before a third booster, those who initially received inactivated vaccines had the lowest rate of positive ELISPOT test (31%), followed by AZD1222/AZD1222 (67%), and AZD1222/BNT162b2 (73%) (p = 0.05), which increased to 90%, 93%, and 100% after the third booster (p=0.4).

### Safety, reactogenicity and disease activity of BNT162b2 booster

The BNT162b2 boosters were well tolerated, and all reactions were mild and transient. The three most common reactions were injection site pain, fatigue and fever (**supplemental figure 3**). Anti-RBD IgG levels did not differ between those without adverse reactions and those who had at least one reaction (p=0.4)

Two SLE patients experienced disease flare after the fourth booster dose. One patient developed rapidly progressive glomerulonephritis from lupus nephritis 1 week after receiving the fourth booster vaccine. Mycophenolate mofetil also had been withheld a few months before the booster administration due to a history of multiple infections. The other developed severe thrombocytopenia 4 weeks after receiving a booster dose. The patient’s disease had been inactive for several years and she had not been receiving immunosuppressive treatment prior to this episode of flare.

## Discussion

As the Omicron variant continues to spread around the world and display increased immune escape potential, aggregating data to optimize immunization strategies in autoimmune rheumatic disease populations is crucial. This study found that a third booster dose of BNT162b2 in SLE and RA patients was well tolerated and elicited a strong humoral immune response against the Omicron variant. However, approximately half of our patients still had a poor response to the third BNT162b2 booster, particularly those who received more intense immunosuppressive treatment or an inactivated vaccine in the initial vaccine series.

By giving a fourth BNT162b2 booster to poor immune responders, anti-RBD IgG increased to a lesser extent compared to the third booster dose (7 fold versus 15 fold), possibly due to the short interval between the third and fourth booster^10^.

Despite the increase of anti-RBD IgG, no discernible increase in neutralizing activity against the Omicron variant was observed. In addition, two SLE patients (7%) experienced severe flares shortly after receiving their fourth booster dose. Multiple studies have reported acceptable safety profiles in SLE patients after initial vaccine series ^11-12^. However, in an animal model, repeated exposure to the same antigen was shown to induce autoimmunity^13^. Given this uncertainty, the benefit of administering a short interval fourth booster dose should be balanced against the risk of disease activation.

In terms of cellular immunity, a third BNT162b2 booster elicited T-cell responses in nearly all of our patients. Positive cellular immune response rates were comparable across initial vaccine types, even among those who received inactivated vaccines, which are known to induce lower a T-cell response ^14^. Recent studies have demonstrated that while variants of concern can evade neutralizing antibodies, the cell-mediated immune response cross-reacted with the variants of concern ^15-18^. It also confers resilient protection against severe disease as antibody titers wane overtime ^19^.

It should be noted that there were several limitations to this study. The findings are observational and based on a small sample size, so they should be interpreted with caution. The results may not be generalizable to other autoimmune rheumatic conditions, other immunosuppressive regimens such as B-cell depletion therapy, or initial mRNA vaccine series. The immunosuppressive load calculation was adapted from the prior study which was arbitrarily determined.

In conclusion, this study demonstrated that the third BNT162b2 booster was well tolerated and significantly improved humoral and cellular immunogenicity in SLE and RA patients. The intensity of immunosuppressive therapy and the type of initial vaccine appear to affect the humoral immune response to the third booster. In poor humoral responders, a fourth BNT162b2 booster administered at a short interval may not provide additional protection against the omicron variant. Our finding that severe flares were observed in SLE patients after the fourth booster is also cause for caution and additional research on the appropriateness of a fourth booster and the timing of such booster in our population.

## Data Availability

All data produced in the present study are available upon reasonable request to the authors

## Acknowledgement

BNT162b2 (lot number 30125BA) used in this study was supplied by the Department of Disease Control, Ministry of Public health with support of Dr Wisit Prasitsirikul, Deputy director of Bamrasnaradura Infectious Disease Institute. The authors thank Mahachakri Sirindhorn Clinical Research Center, Faculty of Medicine, Chulalongkorn University for facility support, and Ms Jariya Pongsaisopon and Sukanlaya Yoosomsuk for coordinating the study. Thanks are also due to Dr Supranee Buranapraditkun and Dr Sasiwimol Ubolyam for enzyme-linked immunosorbent spot testing, and Ms Sutthinee Lapchai for sample preparation and coding. Study data were collected and managed using REDCap electronic data capture tools hosted at Chula Data Management Center - Faculty of Medicine - Chulalongkorn University. We thank Chula Data Management Center, Faculty of Medicine, Chulalongkorn University for the REDCap administrative support.

## Patient and public involvement statement

Patients or the public WERE NOT involved in the design, or conduct, or reporting, or dissemination plans of our research

## Contributors

TA, SS and WK contributed to the conception and design of the work; acquisition, analysis, interpretation of data; and drafting and revising of the work. YP, PV and NW contributed to data acquisition and final approval of the version to be published.

---

**Supplemental figure 1:**
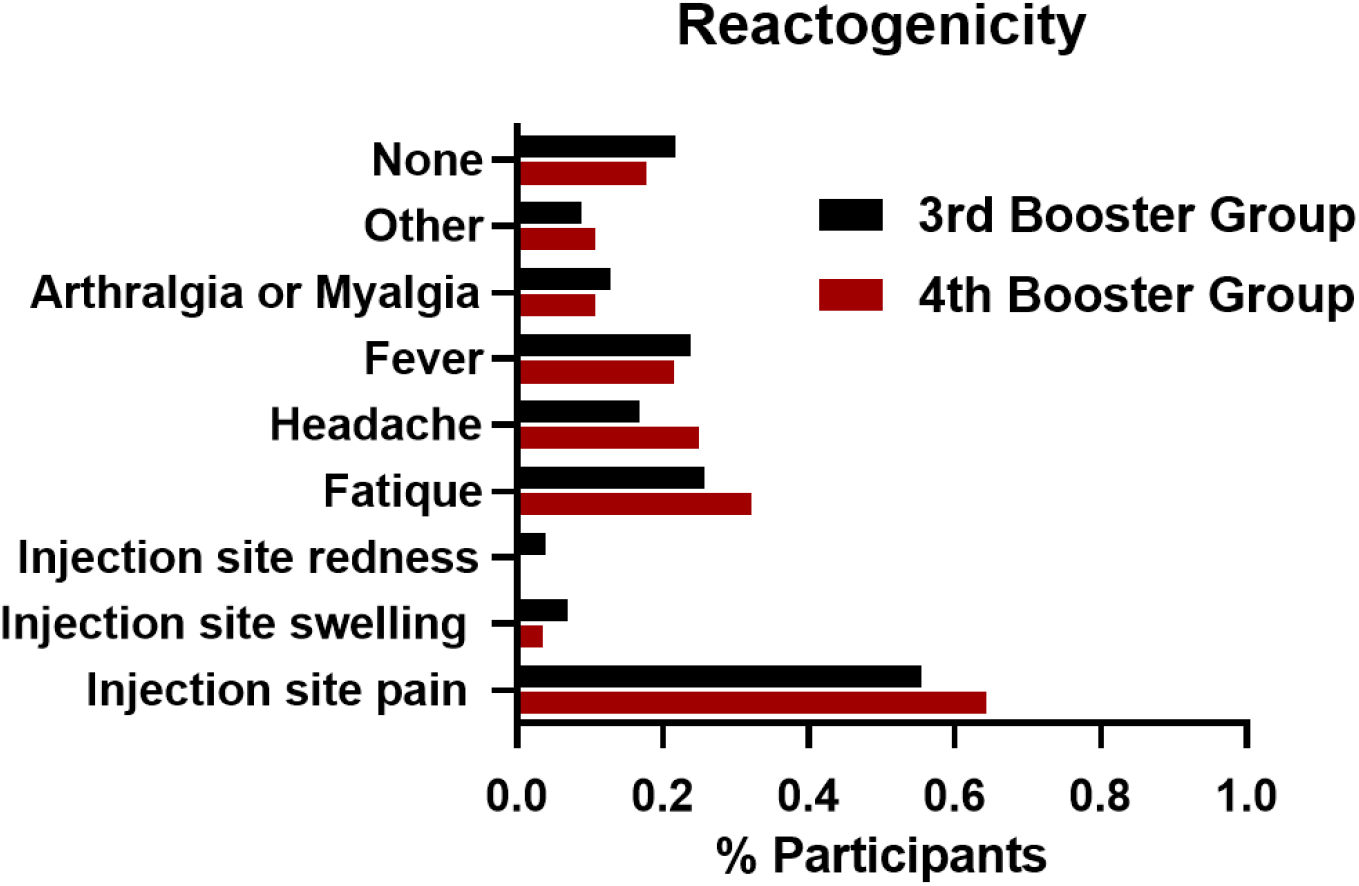
Reactogenicity after the third and fourth BNT162b2 booster doses.

